# Comparison of Automated MRI Perfusion Analysis Software: Agreement in Ischemic Penumbra Estimation and Decision-Making for Endovascular Thrombectomy

**DOI:** 10.1101/2025.08.07.25333241

**Authors:** Jonguk Kim, Jong-Hyeok Park, Dongmin Kim, Myungjae Lee, Joon-Tae Kim, Leonard Sunwoo, Cheolkyu Jung, Wi-Sun Ryu, Beom Joon Kim

## Abstract

**Background:** While computed tomography perfusion is widely used in acute stroke imaging, magnetic resonance perfusion-weighted imaging (PWI) offers superior spatial resolution and tissue specificity, particularly when combined with diffusion-weighted imaging (DWI). However, no prior study has systematically compared automated PWI analysis platforms. This study aims to evaluate the performance of a newly developed software (JLK PWI) against the established RAPID platform in terms of volumetric agreement and clinical decision concordance.

**Methods:** This retrospective multicenter study included 299 patients with acute ischemic stroke who underwent PWI within 24 hours of symptom onset. Volumetric agreement between RAPID and JLK PWI was assessed using concordance correlation coefficients (CCC), Bland–Altman plots, and Pearson correlations. Agreement in endovascular therapy (EVT) eligibility was evaluated using Cohen’s kappa based on DAWN and DEFUSE-3 criteria.

**Results:** The mean age was 70.9 years, 55.9% were male, and the median NIHSS score was 11 (IQR 5–17). The median time from the last known well to PWI was 6.0 hours. JLK PWI showed excellent agreement with RAPID for ischemic core (CCC=0.87; p<0.001) and hypoperfused volume (CCC = 0.88; p<0.001). EVT eligibility classifications based on DAWN criteria showed very high concordance across subgroups (κ=0.85–0.91), and substantial agreement was observed using DEFUSE-3 criteria (κ=0.71).

**Conclusion:** JLK PWI demonstrates high technical and clinical concordance with RAPID, supporting its use as a reliable alternative for MRI-based perfusion analysis in acute stroke care.

## Introduction

The advent of automated perfusion imaging analysis has significantly improved the triage of patients with acute ischemic stroke, particularly by extending the treatment window for endovascular therapy.^1, 2^ Computed tomography perfusion (CTP) has become the predominant modality in emergency settings due to its rapid acquisition and broad accessibility.^3^ As a result, many studies have compared CTP-based software platforms in terms of infarct core estimation, perfusion mismatch, and outcome prediction.^4–6^

In contrast, magnetic resonance perfusion-weighted imaging (PWI) has received far less attention in the context of automated analysis. Although PWI is often used in combination with diffusion-weighted imaging (DWI) in specialized stroke centers, no prior study has systematically compared the outputs of different PWI analysis platforms. This lack of validation has hindered efforts to standardize MRI-based stroke workflows, despite their growing clinical applications.^7, 8^

PWI offers several technical advantages over CTP. It provides higher spatial resolution, is free from beam-hardening artifacts, and is less susceptible to contrast timing errors.^9, 10^ These features improve image quality, particularly in challenging regions such as the posterior fossa or in patients with small vessel disease. Additionally, when paired with DWI, PWI enables more accurate delineation of infarct core and penumbra,^11^ and avoids the risk of radiation exposure,^12^ making it suitable for selected patient populations and research contexts.

Recent clinical trials^13–15^ targeting medium vessel occlusion (MeVO) have underscored the need for more refined imaging biomarkers to better identify patients who may benefit from treatment.^16–18^ The combined spatial precision and tissue specificity of PWI-DWI may enhance patient stratification and inform more personalized treatment strategies.

In this study, we introduce a newly developed PWI analysis platform (JLK PWI, JLK Inc., Republic of Korea) and compare its performance with that of a widely used commercial software (RAPID, RAPID AI, CA, USA). We evaluate inter-platform agreement in volumetric parameters, including ischemic core, hypoperfused area, and mismatch volume, as well as in treatment eligibility based on DAWN^1^ and DEFUSE-3^2^ trial criteria. This study aims to evaluate the clinical viability of JLK PWI as a robust alternative for MRI-based stroke assessment.

## Methods

### Study Design and Study Population

This retrospective multicenter study included patients with acute ischemic stroke who underwent PWI within 24 hours of symptom onset at two tertiary hospitals in Korea. A total of 216 patients from Seoul National University Bundang Hospital who underwent both PWI and endovascular thrombectomy between January 2019 and April 2024, and 102 patients from Chonnam National University Hospital who underwent PWI within 24 hours of symptom onset with or without endovascular thrombectomy (EVT) between January 2015 and December 2015, were initially screened. After pooling the datasets, 318 patients met the inclusion criteria. Of these, patients were excluded due to abnormal arterial input function (n=6), severe motion artifacts (n=2), or inadequate image (n=11). Consequently, 299 patients were included in the final analysis. The study protocol was approved by the institutional review board of Seoul National University Bundang hospital [IRB# B-1710-429-102], and written informed consent was obtained from all patients or their legal representatives.

### Clinical Data Collection

Using a standardized protocol,^19^ we prospectively collected demographic data, vascular risk factors (hypertension, diabetes mellitus, hyperlipidemia, coronary artery disease, atrial fibrillation, and smoking history), prior medication use, pre-stroke functional status, and index stroke characteristics, such as initial stroke severity (NIH Stroke Scale, NIHSS) and subtypes. Stroke subtypes were determined by an experienced vascular neurologist, using a validated MRI-based classification system built on the TOAST (Trial of ORG 10172 in Acute Stroke Treatment) criteria.^20^

### Imaging and Image Reconstruction

All perfusion MRI scans were performed on either 3.0T (62.3%) or 1.5T (37.7%) scanners. Regarding the vendors, 32.4% of scans were conducted using GE systems, 61.9% using Philips systems, and 5.7% using Siemens systems, all equipped with an 8-channel head coil. Dynamic susceptibility contrast-enhanced perfusion imaging was performed using a gradient-echo echo-planar imaging (GE-EPI) sequence. The imaging parameters were as follows: repetition time (TR) = 1,000-1,500 ms (6.3%), 1,500-2,000 ms (66.7%), or 2,000-2,500 ms (27.0%); echo time (TE) = 30-40 ms (1.0%), 40-50 ms (91.8%), or 60-70 (7.2%); field of view (FOV) = 210×210 mm^2^ (5.7%), or 230×230 mm^2^ (94.3%); and slice thickness of 5 mm with no interslice gap, covering the entire supratentorial brain with 17–25 slices. Images were reconstructed and exported in DICOM format for subsequent post-processing and quantitative perfusion analysis. All image analyses were done in the central image laboratory operated by Seoul National University Bundang Hospital.

### Automated PWI Analysis

For infarct core estimation, RAPID employed the default threshold of ADC < 620 × 10^−^□ mm^2^/s. JLK PWI utilized a validated deep learning–based infarct segmentation algorithm applied to the b1000 DWI images.^21–23^ For delineating the hypoperfused region, both platforms used the conventional threshold of T_max_ > 6 seconds. Preprocessing steps including motion correction, skull stripping, vessel masking, and perfusion parameter calculations were performed automatically by each platform. All segmentations and resulting images were visually inspected to ensure technical adequacy before inclusion in the analysis.

### Statistical Analysis

Descriptive statistics were used to summarize baseline characteristics. Continuous variables were reported as means with standard deviations (SD) or medians with interquartile ranges (IQR), depending on data distribution. Categorical variables were presented as counts with percentages. Agreement between the two platforms in perfusion parameter measurements (ischemic core volume, hypoperfused volume, and mismatch volume) was assessed using concordance correlation coefficients (CCC), Pearson correlation coefficients, and Bland–Altman plots. The magnitude of agreement was classified as: poor (0.0–0.2), fair (0.21–0.40), moderate (0.41–0.60), substantial (0.61–0.80), and excellent (0.81–1.0).

For EVT eligibility, classification agreement between the RAPID and JLK software was evaluated using Cohen’s kappa coefficient, applied separately for each subgroup defined by the DAWN and DEFUSE-3 trial criteria. The DAWN classification stratified eligible infarct volume based on age and NIHSS into three prespecified categories, while the DEFUSE-3 classification used a mismatch ratio ≥1.8 and an infarct core volume <70mL. Cases with discordant EVT eligibility classifications were additionally analyzed descriptively. Subgroup analyses were conducted for patients with anterior circulation large vessel occlusion and those with basilar artery occlusion. In each subgroup, agreement metrics and outcome prediction models were separately generated to evaluate software performance across stroke types.

All statistical analyses were performed using STATA version 16.0 (StataCorp LLC, College Station, TX) and R version 4.2.3 (R Foundation for Statistical Computing, Vienna, Austria). A two-sided p-value < 0.05 was considered statistically significant.

## Results

### Subject Characteristics

For 299 subjects included, the mean age was 70.9 years (SD 11.6), and 55.9% were male. The median NIHSS score on admission was 11 (IQR: 5–17). The most common stroke subtype was cardioembolism (45.2%), followed by large artery atherosclerosis (29.1%) and undetermined etiology (13.0%). Intravenous thrombolysis was administered in 157 patients (52.5%).

Regarding occlusion sites, 208 (69.6%) subjects had anterior circulation large vessel occlusion, and 31 had basilar artery occlusion (10.4%). Meanwhile, 60 (20.1%) subjects had no large vessel occlusion on MRI. The median time from the last known well to PWI was 360 (IQR: 216 – 750) minues6.0 hours (IQR: 3.6-12.5), and the median time from PWI to groin puncture was 55.5 minutes (IRQ: 40.8-82.3).

### Concordance of Ischemic Core, Hypoperfused, and Mismatch Volumes

Ischemic core volumes showed excellent agreement between RAPID and JLK PWI, with CCC = 0.87 (95% CI, 0.77 – 0.94; Figure 1B). The Bland-Altman plot showed a mean difference of −4.05 mL and limits of agreement ranging from −41.62 to 33.53 mL (Figure 1A). Similarly, hypoperfused volumes showed excellent agreement (CCC = 0.88 [95% CI, 0.80 – 0.93]; Figure 1D). The mean difference was 2.46 mL, with limits of agreement from −59.37 to 64.30 mL (Figure 1C). Mismatch volumes demonstrated substantial agreement (CCC = 0.78 [95% CI, 0.69 – 0.84]; Figure 1F), with a mean difference of 6.51 mL and limits of agreement from −68.86 to 81.88 mL (Figure 1E).

**Figure 1.**
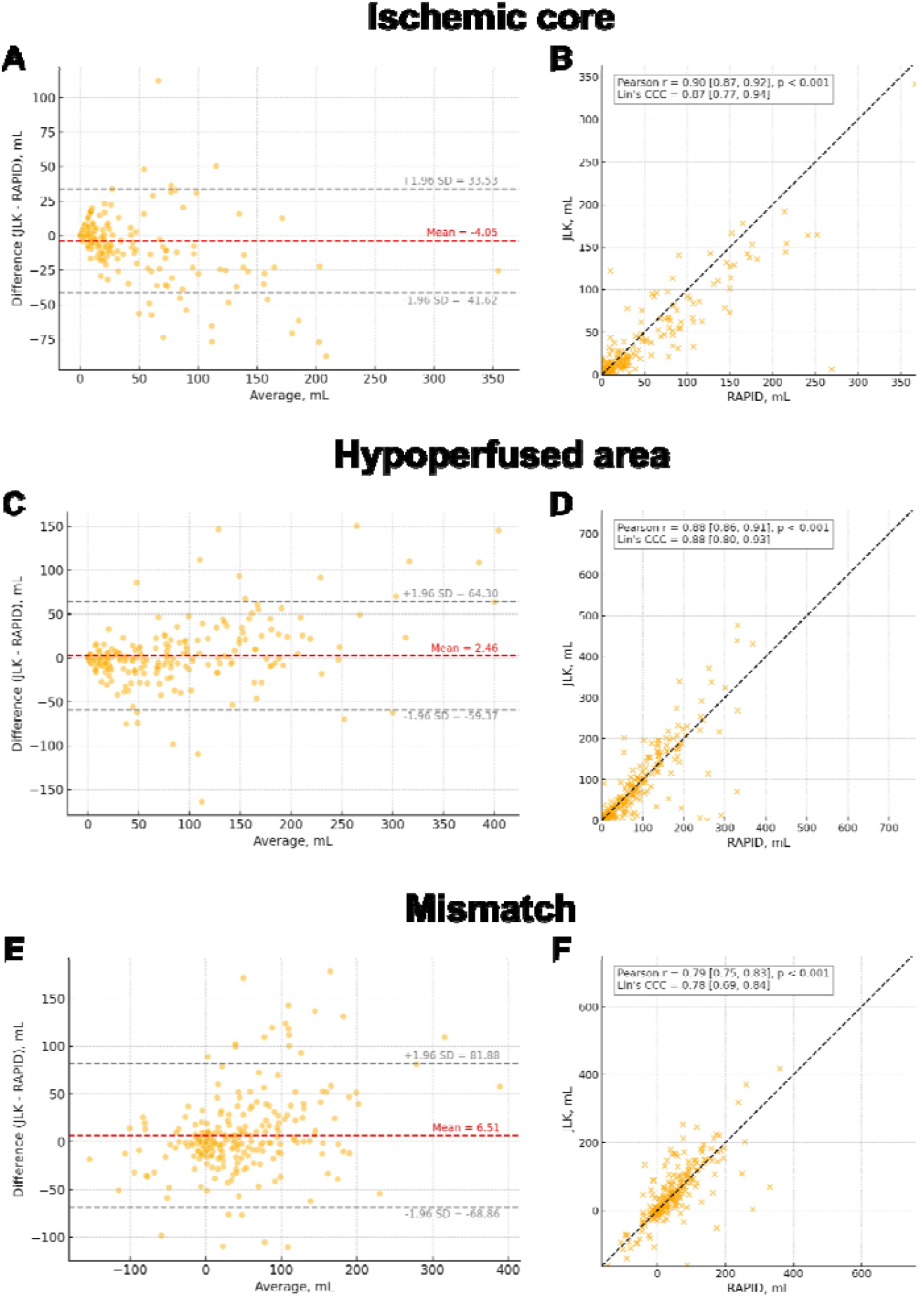
Concordance of Ischemic Core, Hypoperfused Area, and Mismatch Volumes Between Platforms. (A) Bland–Altman plot and (B) scatter plot fo ischemic core volumes. (C) Bland–Altman plot and (D) scatter plot for hypoperfused volumes. (E) Bland–Altman plot and (F) scatter plot for mismatch volumes. Red dotted lines indicate the mean difference, and gray dotted lines represent the limits of agreement between the two platforms.

Subgroup analyses for patients with anterior circulation large vessel occlusion (Supplementary Figure 1) and basilar artery occlusion (Supplementary Figure 2) showed similarly trends in agreement across core, hypoperfusion, and mismatch volumes.

**Figure 2.**
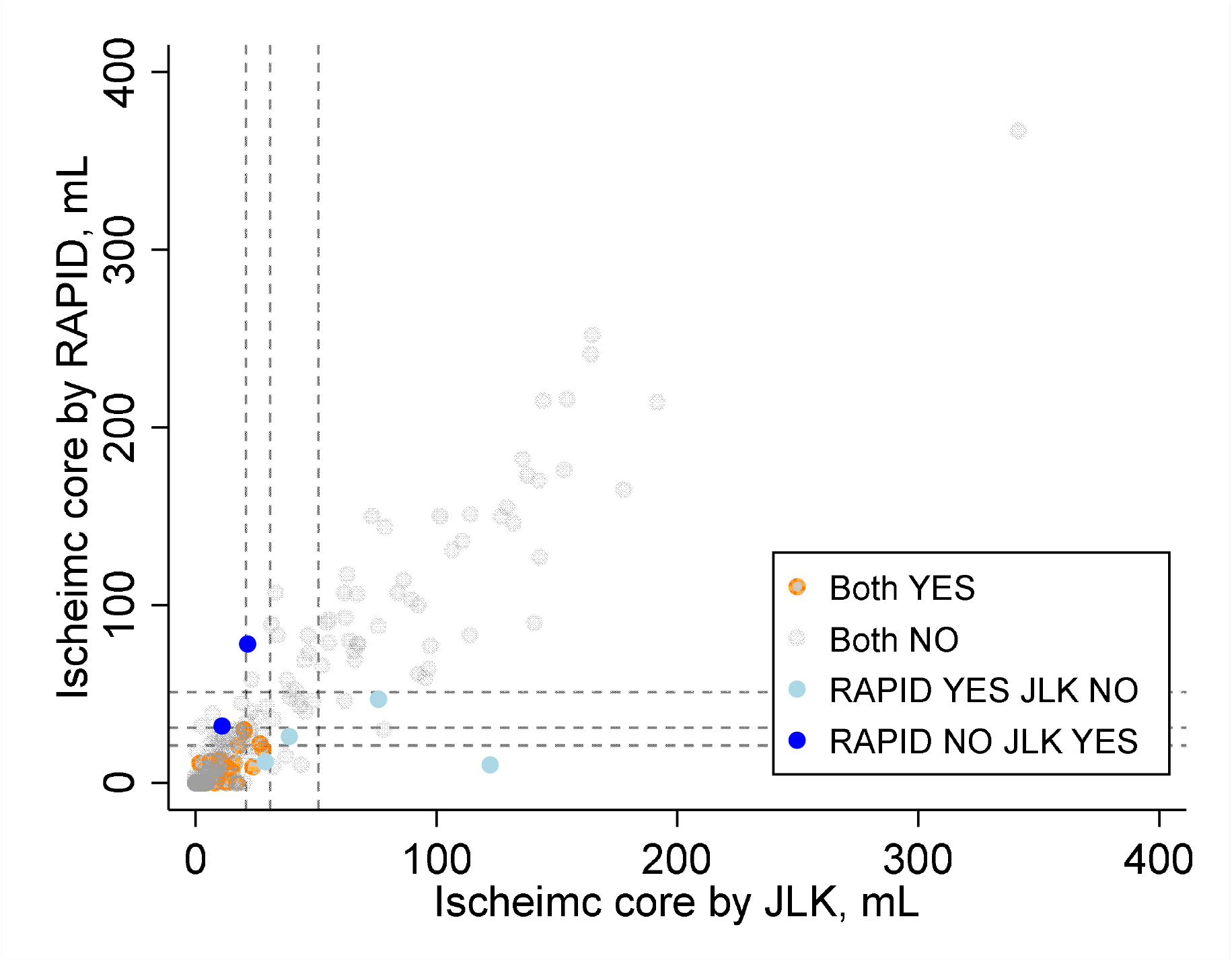
Comparison of RAPID and JLK Software for Endovascular Thrombectomy Eligibility Based on Variable Core Volume Criteria from the DAWN Trial. Gray dotted lines indicate core volume of 21, 31, and 50 mL

### Concordance of EVT Eligibility Based on DAWN and DEFUSE-3 Criteria

When applying the DAWN trial’s criteria, agreement between RAPID and JLK PWI in determining EVT eligibility was excellent, with an overall Cohen’s κ values of 0.895 (95% CI, 0.812 - 0.978). Agreement remained high across all subgroups defined by age and NIHSS score: 0.852 in patients aged >80 years with NIHSS ≥10 (n=51; Table 2; Figure 2), 0.904 in those aged ≤80 with NIHSS 10–19 (n=100), and 0.857 in those aged ≤80 with NIHSS ≥20 (n=20).

**Table 1.**
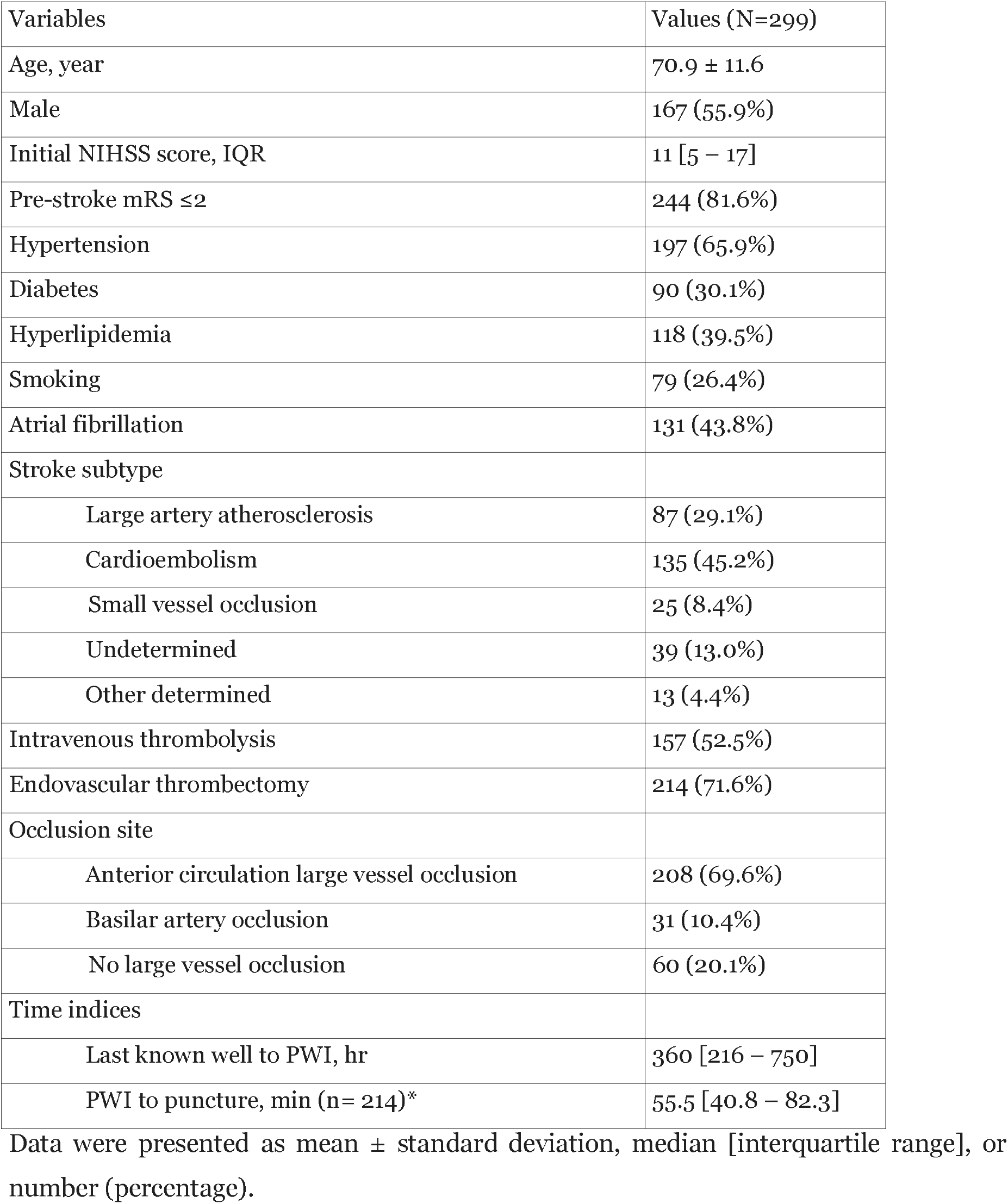
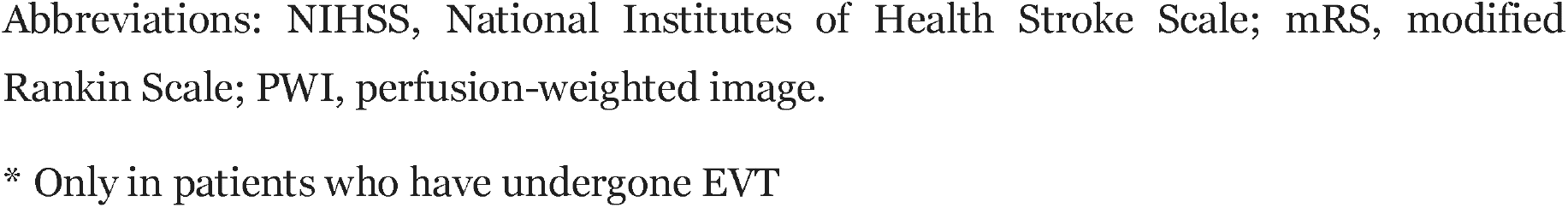
Baseline Characteristics of the Study Population.

**Table 2.**
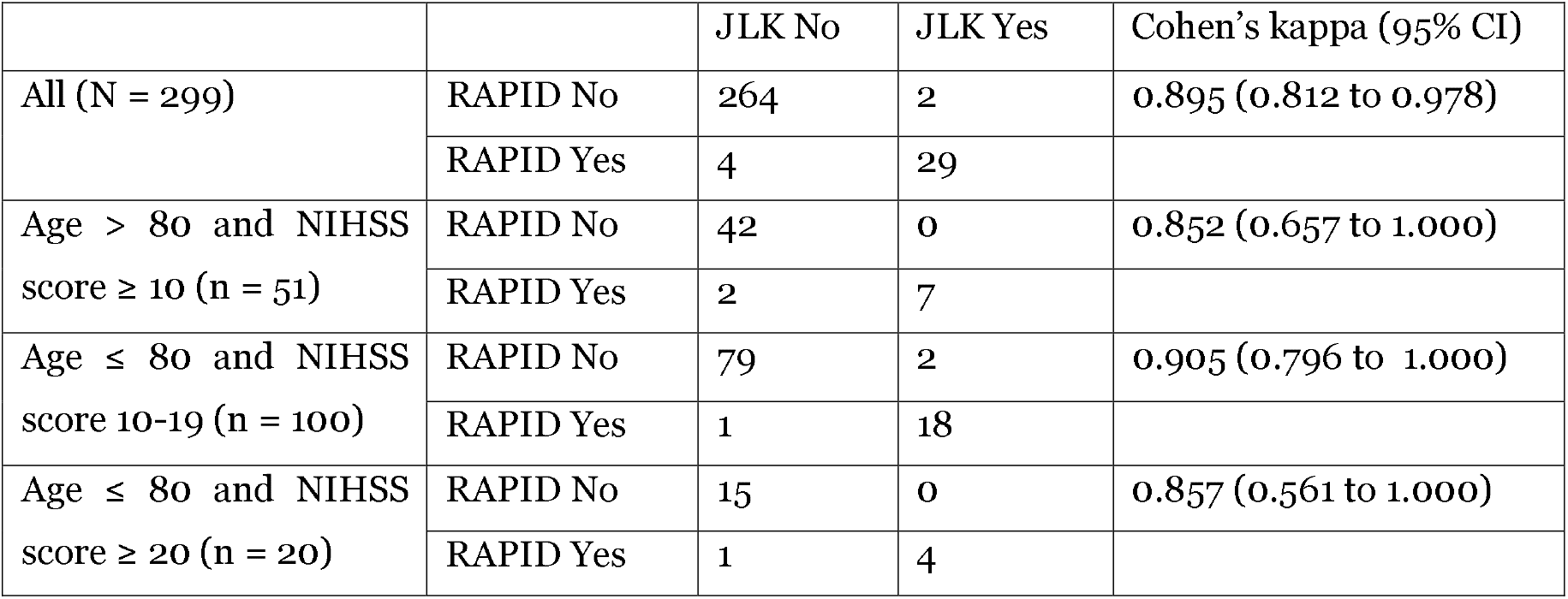
Assessment of RAPID vs. JLK Software in Determining Eligibility for Endovascular Thrombectomy Based on DAWN Trial Criteria.

When using the DEFUSE-3 trial’s criteria, the agreement was substantial (Cohen’s κ of 0.706). Both platforms labeled 125 patients as EVT-eligible and 82 as ineligible. However, 39 patients were discordantly classified (17 eligible by RAPID only, 22 by JLK PWI only, Figure 3).

**Figure 3.**
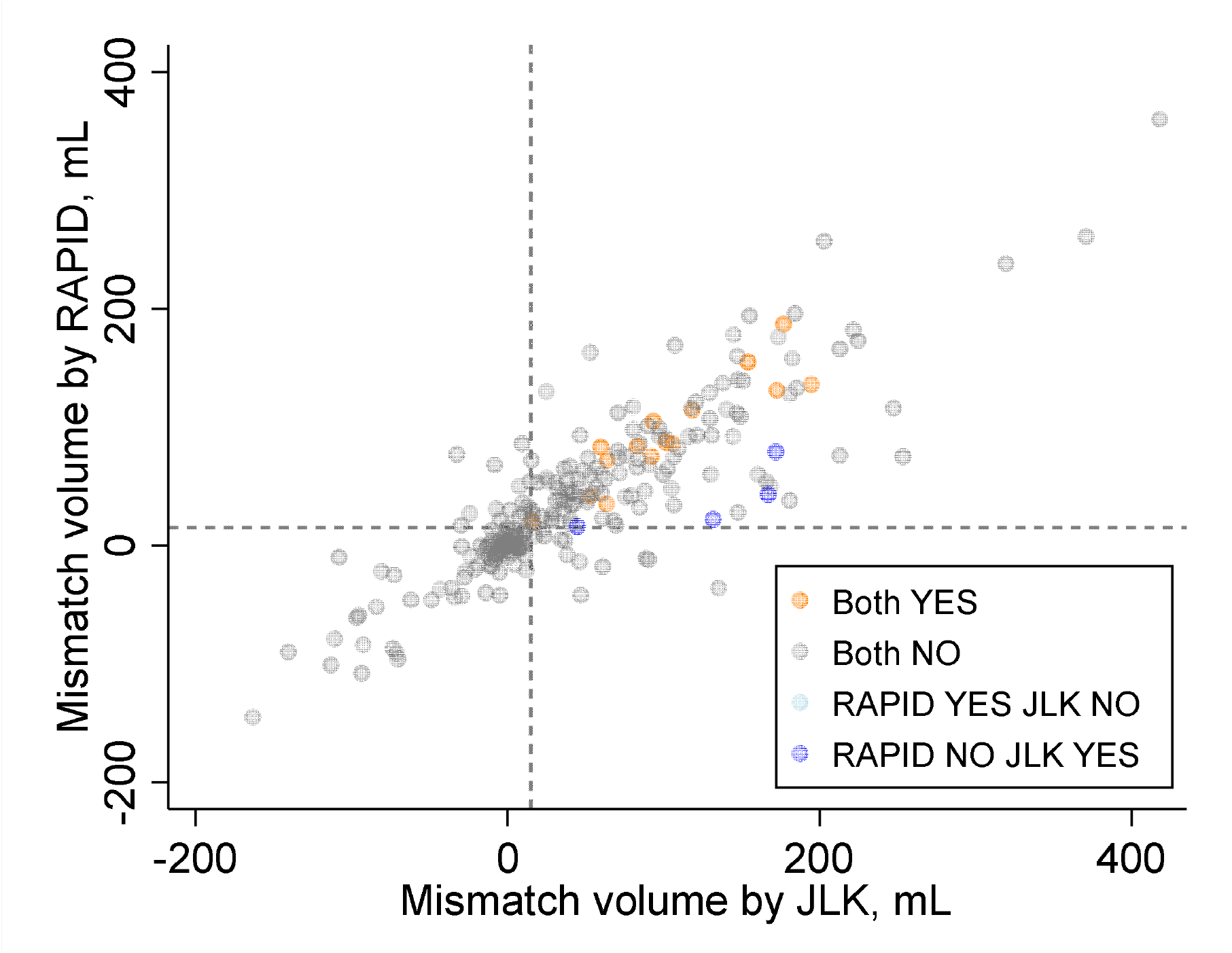
Comparison of RAPID and JLK Software for EVT Eligibility According to Mismatch Volume and Ratio Thresholds Defined in the DEFUSE 3 Trial. Gray dotted lines indicate mismatch volume of 15 mL.

## Discussion

To our knowledge, this study is among the first to conduct a comprehensive validation of a newly developed MRI perfusion software (JLK PWI) against the established RAPID platform, using both volumetric and clinical decision-making metrics. Importantly, our analysis is not limited to core–hypoperfusion volume comparisons but also includes EVT triage concordance based on DAWN and DEFUSE-3 criteria, as well as segmentation alignment with diffusion-restricted lesions (ADC < 620). This multifaceted approach offers a pragmatic perspective for assessing real-world performance of automated perfusion software within acute stroke workflows.

A notable strength of our study lies in its inclusion of broad stroke population, encompassing both anterior and posterior circulation large vessel occlusion, and a wide spectrum of imaging time windows up to 24 hours. Most prior validation studies have focused on CTP-derived perfusion maps or DWI-based core estimation alone.^4, 24^ By leveraging PWI-DWI integration in a clinical setting, we demonstrate that the JLK PWI achieves excellent volumetric agreement with RAPID (CCC = 0.87) and high agreement in EVT decision-making (Cohen’s κ up to 0.90 for DAWN). These results support the use of JLK PWI not only as a technical substitute, but also as a clinical decision-making tool.^25, 26^

Accurate, automated estimation of infarct core is critical for patient selection in reperfusion therapies, particularly in extended time windows and in settings where CTP is unavailable or unsuitable.^25, 27^ Our findings show that JLK PWI maintains high fidelity in infarct core and hypoperfusion volume estimation across diverse patient profiles. Notably, EVT eligibility classifications showed over 85% agreement using both DAWN and DEFUSE-3 frameworks, suggesting that JLK PWI can be effective in guideline-based treatment decisions.

Furthermore, JLK PWI software demonstrated robust performance even in posterior circulation strokes, where perfusion analysis remains technically challenging.^28^ This supports its potential utility in future studies and clinical protocols involving basilar occlusions or MeVOs, which are increasingly recognized as important therapeutic targets despite the current lack of standardized imaging criteria.^29, 30^

Some limitations must be acknowledged. First, the generalizability of our findings may be limited by the retrospective design and the inclusion of two tertiary stroke centers. Second, although both hospitals used 3T MRI systems, subtle differences in acquisition parameters and contrast timing may influence software outputs. Third, we did not include specific imaging data such as infarct growth or collateral status,^31^ which could provide additional context for discrepancies in EVT decision classification.

Future studies should prospectively validate JLK PWI in broader clinical settings, including underrepresented stroke populations such as MeVOs and wake-up strokes. Integration of additional imaging biomarkers, such as collateral grading,^32^ or radiomic texture features,^33^ may further enhance prediction models and assist in complex clinical decisions. In addition, longitudinal studies examining infarct evolution and clinical outcomes after EVT could provide insight into the long-term predictive validity of automated core estimation tools like JLK PWI.

## Conclusion

In conclusion, JLK PWI demonstrates excellent technical and clinical agreement with established commercial software, offering reliable infarct core estimation and high concordance in EVT decision-making. Its applicability across diverse stroke types and its foundation in deep learning–based analysis position it as a promising tool for real-time stroke triage. As imaging-based selection becomes increasingly nuanced, JLK PWI may play a critical role in improving access to individualized, time-sensitive reperfusion therapy.

## Data Availability

All data produced in the present study are available upon reasonable request to the authors.

## Acknowledgment

The authors appreciate the contributions of all members of the Clinical Research Collaboration for Stroke in Korea to this study.

## Supplemental Material

Supplementary Figure 1.

Supplementary Figure 2.

## References

1. Jovin TG, Nogueira RG, Investigators D. Thrombectomy 6 to 24 Hours after Stroke. N Engl J Med. 2018;378(12):1161–1162.

2. Albers GW, Marks MP, Kemp S, et al. Thrombectomy for Stroke at 6 to 16 Hours with Selection by Perfusion Imaging. N Engl J Med. 2018;378(8):708–718.

3. Kim N, Ryu WS, Ha SY, et al. Optimal Cerebral Blood Flow Thresholds for Ischemic Core Estimation Using Computed Tomography Perfusion and Diffusion-Weighted Imaging. Ann Neurol. 2025;97(5):919–929.

4. Kim N, Ha SY, Park GH, et al. Comparison of two automated CT perfusion software packages in patients with ischemic stroke presenting within 24 h of onset. Front Neurosci. 2024;18:1398889.

5. Suomalainen OP, Martinez-Majander N, Sibolt G, et al. Comparative analysis of core and perfusion lesion volumes between commercially available computed tomography perfusion software. Eur Stroke J. 2023;8(1):259–267.

6. Xiong Y, Huang CC, Fisher M, Hackney DB, Bhadelia RA, Selim MH. Comparison of Automated CT Perfusion Softwares in Evaluation of Acute Ischemic Stroke. J Stroke Cerebrovasc Dis. 2019;28(12):104392.

7. Xiong Y, Luo Y, Wang M, et al. Evaluation of Diffusion-Perfusion Mismatch in Acute Ischemic Stroke with a New Automated Perfusion-Weighted Imaging Software: A Retrospective Study. Neurol Ther. 2022;11(4):1777–1788.

8. Chatterjee NR, Ansari SA, Vakil P, Prabhakaran S, Carroll TJ, Hurley MC. Automated analysis of perfusion weighted MRI using asymmetry in vascular territories. Magn Reson Imaging. 2015;33(5):618–23.

9. Konstas AA, Goldmakher GV, Lee TY, Lev MH. Theoretic basis and technical implementations of CT perfusion in acute ischemic stroke, part 1: Theoretic basis. AJNR Am J Neuroradiol. 2009;30(4):662–8.

10. Liu M, Wen X, Li M, et al. Blind spots in brain imaging: a pictorial essay. Quant Imaging Med Surg. 2025;15(1):1023–1039.

11. Kane I, Sandercock P, Wardlaw J. Magnetic resonance perfusion diffusion mismatch and thrombolysis in acute ischaemic stroke: a systematic review of the evidence to date. J Neurol Neurosurg Psychiatry. 2007;78(5):485–91.

12. Cohnen M, Wittsack HJ, Assadi S, et al. Radiation exposure of patients in comprehensive computed tomography of the head in acute stroke. AJNR Am J Neuroradiol. 2006;27(8):1741–5.

13. Psychogios M, Brehm A, Ribo M, et al. Endovascular Treatment for Stroke Due to Occlusion of Medium or Distal Vessels. N Engl J Med. 2025;392(14):1374–1384.

14. Goyal M, Ospel JM, Ganesh A, et al. Endovascular Treatment of Stroke Due to Medium-Vessel Occlusion. N Engl J Med. 2025;392(14):1385–1395.

15. Mohammaden MH, Souza Viana L, Abdelhamid H, et al. Endovascular Versus Medical Management in Distal Medium Vessel Occlusion Stroke: The DUSK Study. Stroke. 2024;55(6):1489–1497.

16. Ospel JM, Nguyen TN, Jadhav AP, et al. Endovascular Treatment of Medium Vessel Occlusion Stroke. Stroke. 2024;55(3):769–778.

17. Salim HA, Vagal V, Lakhani DA, et al. Association of Pretreatment Perfusion Imaging Parameters with 90-Day Excellent Functional Outcomes in Anterior Circulation Distal Medium Vessel Occlusion Stroke. AJNR Am J Neuroradiol. 2025;46(5):892–899.

18. Cai LY, Hoseinyazdi M, Lakhani DA, et al. Redefining ischemic core, penumbra, and target mismatch on CT perfusion in acute anterior distal medium vessel occlusion. medRxiv. 2025:2025.03.25.25324574.

19. Kim BJ, Park JM, Kang K, et al. Case characteristics, hyperacute treatment, and outcome information from the clinical research center for stroke-fifth division registry in South Korea. J Stroke. 2015;17(1):38–53.

20. Ko Y, Lee S, Chung J-W, et al. MRI-based algorithm for acute ischemic stroke subtype classification. Journal of stroke. 2014;16(3):161.

21. Ryu WS, Schellingerhout D, Park J, et al. Deep learning-based automatic segmentation of cerebral infarcts on diffusion MRI. Sci Rep. 2025;15(1):13214.

22. Ryu WS, Schellingerhout D, Lee H, et al. Deep Learning-Based Automatic Classification of Ischemic Stroke Subtype Using Diffusion-Weighted Images. J Stroke. 2024;26(2):300–311.

23. Ryu WS, Kang YR, Noh YG, et al. Acute Infarct Segmentation on Diffusion-Weighted Imaging Using Deep Learning Algorithm and RAPID MRI. J Stroke. 2023;25(3):425–429.

24. Austein F, Riedel C, Kerby T, et al. Comparison of Perfusion CT Software to Predict the Final Infarct Volume After Thrombectomy. Stroke. 2016;47(9):2311–7.

25. Mishra NK, Albers GW, Christensen S, et al. Comparison of magnetic resonance imaging mismatch criteria to select patients for endovascular stroke therapy. Stroke. 2014;45(5):1369–74.

26. Neumann-Haefelin T, Wittsack HJ, Wenserski F, et al. Diffusion- and perfusion-weighted MRI. The DWI/PWI mismatch region in acute stroke. Stroke. 1999;30(8):1591–7.

27. Evans JW, Graham BR, Pordeli P, et al. Time for a Time Window Extension: Insights from Late Presenters in the ESCAPE Trial. AJNR Am J Neuroradiol. 2018;39(1):102–106.

28. Pallesen LP, Lambrou D, Eskandari A, et al. Perfusion computed tomography in posterior circulation stroke: predictors and prognostic implications of focal hypoperfusion. Eur J Neurol. 2018;25(5):725–731.

29. Cimflova P, McDonough R, Kappelhof M, et al. Perceived Limits of Endovascular Treatment for Secondary Medium-Vessel-Occlusion Stroke. AJNR Am J Neuroradiol. 2021;42(12):2188–2193.

30. Alemseged F, Nguyen TN, Alverne FM, Liu X, Schonewille WJ, Nogueira RG. Endovascular Therapy for Basilar Artery Occlusion. Stroke. 2023;54(4):1127–1137.

31. de Havenon A, Mlynash M, Kim-Tenser MA, et al. Results From DEFUSE 3: Good Collaterals Are Associated With Reduced Ischemic Core Growth but Not Neurologic Outcome. Stroke. 2019;50(3):632–638.

32. Tetteh G, Navarro F, Meier R, et al. A deep learning approach to predict collateral flow in stroke patients using radiomic features from perfusion images. Front Neurol. 2023;14:1039693.

33. Li M, Jiang J, Gu H, Hu S, Wang J, Hu C. CT-Based Intrathrombus and Perithrombus Radiomics for Prediction of Prognosis after Endovascular Thrombectomy: A Retrospective Study across 2 Centers. AJNR Am J Neuroradiol. 2025;46(4):681–688.

