## Supplementary Material for "Comparison of Automated MRI Perfusion Analysis Software: Agreement in Ischemic Penumbra Estimation and Decision-Making for Endovascular Thrombectomy"

Supplementary Figure 1. Concordance of Ischemic Core, Hypoperfused Area, and Mismatch Volumes Between Platforms in Patients with Anterior Circulation Large Vessel Occlusion.

Supplementary Figure 2. Concordance of Ischemic core, Hypoperfused Area, and Mismatch Volumes Between Platforms in patients with Basilar Artery Occlusion.

**Supplementary Figure 1. Concordance of Ischemic Core, Hypoperfused Area, and Mismatch Volumes Between Platforms in Patients with Anterior Circulation Large Vessel Occlusion.** (A) Bland–Altman plot and (B) scatter plot for ischemic core volumes. (C) Bland–Altman plot and (D) scatter plot for hypoperfused volumes. (E) Bland–Altman plot and (F) scatter plot for mismatch volumes. Red dotted lines indicate the mean difference, and gray dotted lines represent the limits of agreement between the two platforms.

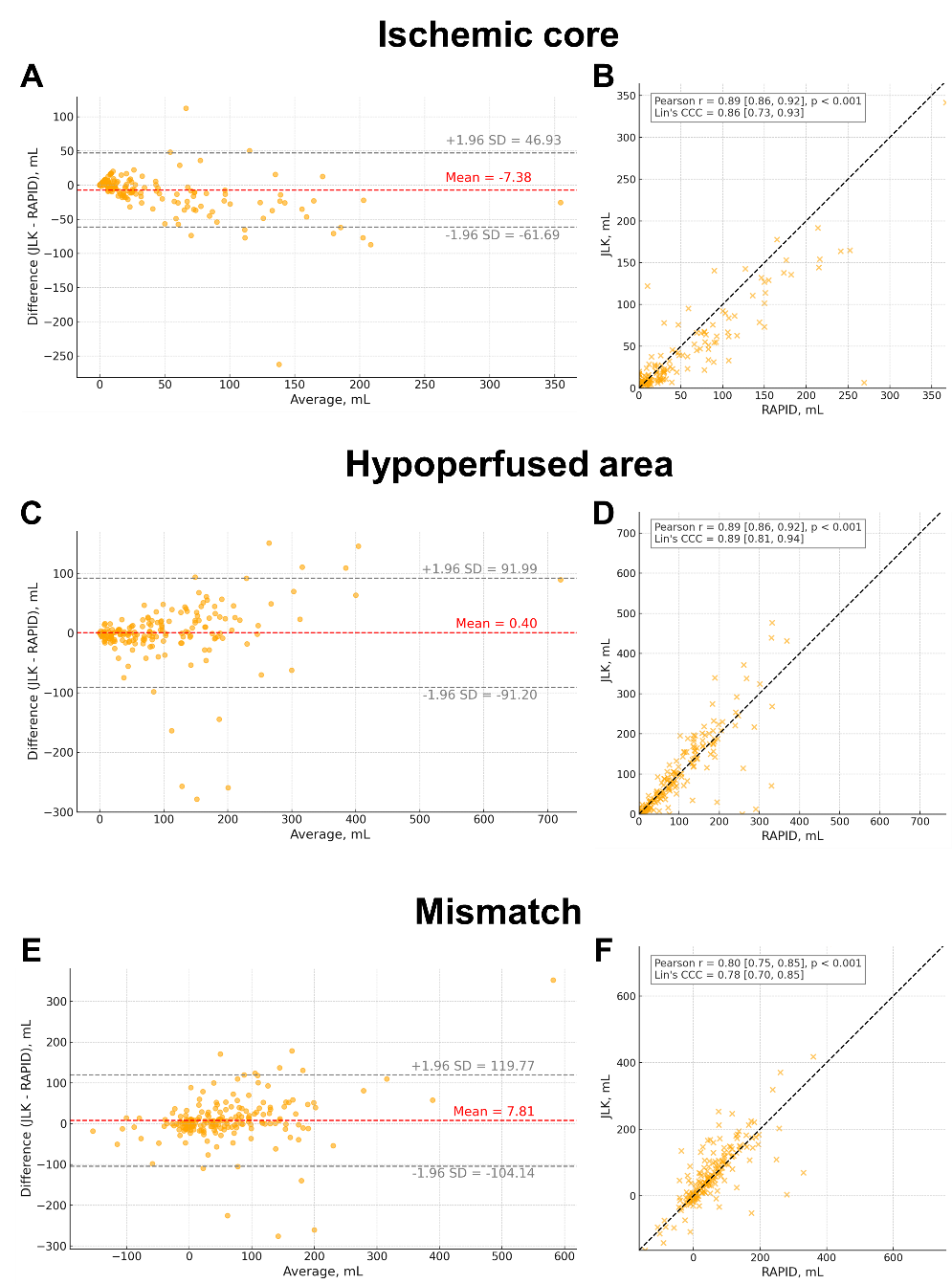

**Supplementary Figure 2. Concordance of Ischemic core, Hypoperfused Area, and Mismatch Volumes Between Platforms in patients with Basilar Artery Occlusion.** (A) Bland–Altman plot and (B) scatter plot for ischemic core volumes. (C) Bland–Altman plot and (D) scatter plot for hypoperfused volumes. (E) Bland–Altman plot and (F) scatter plot for mismatch volumes. Red dotted lines indicate the mean difference, and gray dotted lines represent the limits of agreement between the two platforms.

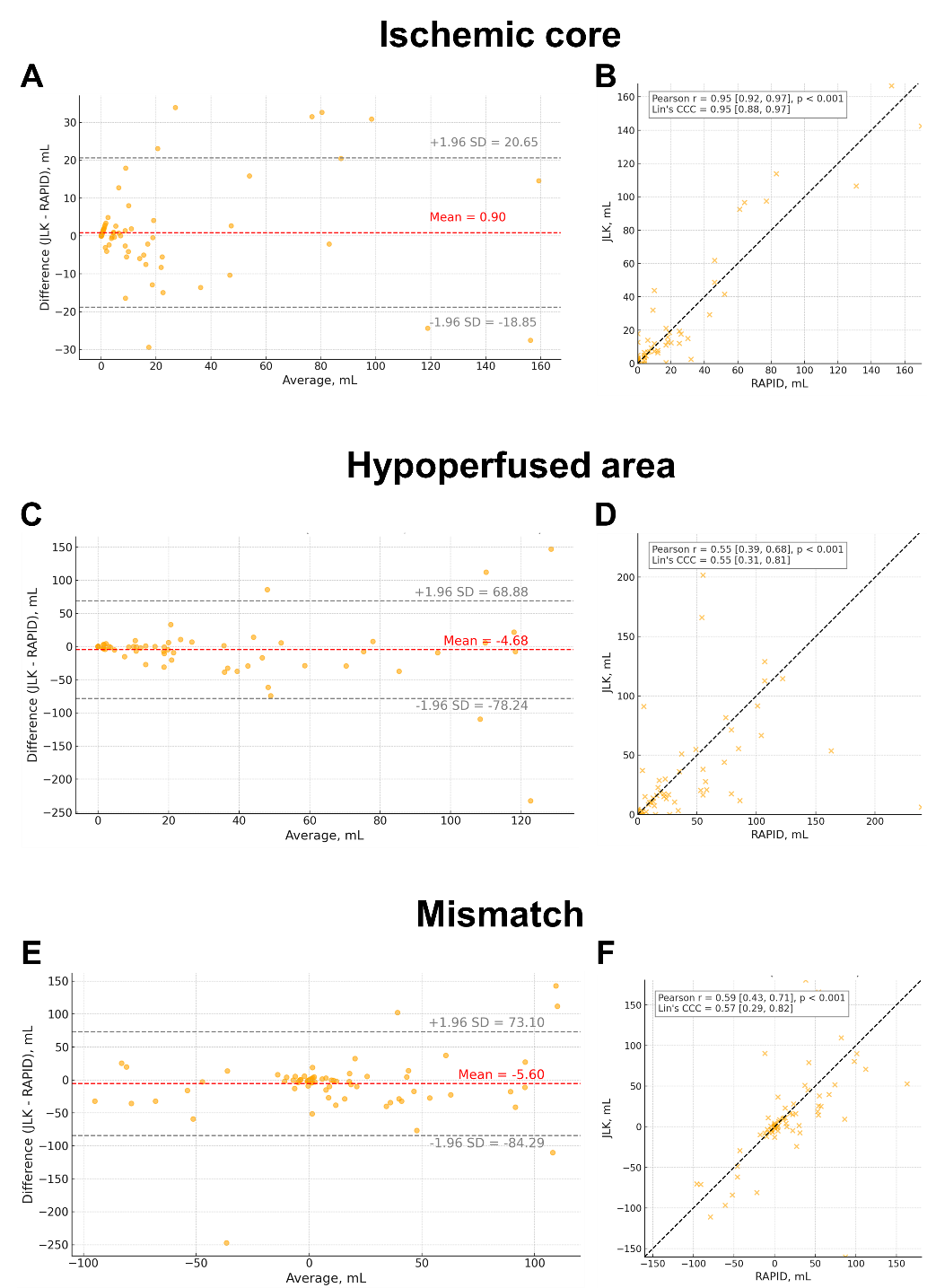
